# Order-Based Bayesian Network Modeling of Early Detection and Post-Diagnosis Control for Cardiovascular Disease Risk in Type 2 Diabetes

**DOI:** 10.64898/2026.06.10.26355419

**Authors:** Yukti Kathuria, Kristen Miller, Elizabeth Selden, William Gallagher, Muge Capan

**Author notes:** These authors contributed equally to this work (Yukti Kathuria, Muge Capan). These authors also contributed equally to this work (Kristen Miller, Elizabeth Selden, William Gallagher).

## Abstract

Patients diagnosed with type 2 diabetes (T2D) are at increased risk of developing cardiovascular disease (CVD), the leading cause of morbidity and mortality in this population. Early detection and glycemic control within the first year after diagnosis reduce CVD risk. However, gaps remain in how to operationalize early detection of T2D using Electronic Health Record (EHR) data and quantify its relationship with subsequent CVD risk using longitudinal observations. We developed a probabilistic graph model to analyze the interdependencies between early detection of T2D, post-diagnosis glycemic control, and CVD occurrence. Using a temporally structured Bayesian Network (BN) learned from EHR data of 9,450 primary care patients between 2017 and 2023, we quantified probabilistic dependencies between demographics, diagnostic delay surrogates, glycemic control, and post-diagnosis CVD occurrence. Percentile-based thresholds defined risk groups, where individuals with predicted probabilities in the bottom decile (≤ 10*^th^* percentile) were classified as low risk, and those in the top decile (≥ 90*^th^* percentile) as high risk. Results demonstrated heterogeneity in predicted risks across glycemic and cardiovascular outcomes. Predicted probability of developing CVD within the first year after T2D diagnosis ranged from a mean of 5.2% in the low-risk group to 28.9% in the high-risk group, while predicted probabilities of mean Hemoglobin A1c (HbA1c) ≥ 8% during the first year post-diagnosis ranged from 1.6% in low-risk to 55.1% in high-risk group. Patients with HbA1c at diagnosis ≥ 8% had higher predicted probabilities of first-year post-diagnosis mean HbA1c ≥ 8% (53.3% vs. 1.9%) and high HbA1c coefficient of variation (18.7% vs. 3.1%) compared with those with HbA1c ≤ 6.5%. Incorporating early clinical outcomes refined later risk predictions, with long-term CVD risk reaching 33.5% among high-risk individuals. The proposed model achieved predictive performance comparable to conventional machine learning approaches while providing interpretable relationships for risk stratification in primary care populations.

**Author summary:** People with type 2 diabetes (T2D) are more likely to develop cardiovascular disease (CVD) after becoming diabetic. Having multiple diseases leads to more illness burden and death. Detecting T2D early and managing its symptoms soon after diagnosis can help prevent CVD, but it is difficult to identify these symptoms in data and understand how they relate to each other. In this study, we used healthcare data from primary care patients to explore how early signs of T2D (such as laboratory measurements), and health complications are connected. We developed a data-driven model that connects patient characteristics, clinical observations, and health outcomes. Our findings showed having high blood sugar values at diagnosis worsened how diabetes is controlled later on and put individuals in higher risk of CVD. Particularly, clinical observations during the first year after T2D diagnosis were important to detect if someone will develop CVD later. By connecting data to health outcomes over time, our model may help clinicians identify high-risk individuals earlier and support more personalized T2D management in primary care settings.

## 1 Introduction

Type 2 diabetes (T2D) is a multi-organ chronic disease that accelerates the development of other conditions, including cardiovascular disease (CVD). CVD is one of the leading causes of morbidity and mortality in T2D population (1). Individuals with T2D experience an increased lifetime risk of cardiovascular complications compared with the general population (2). Evidence suggests that diabetic patients have a two to four times higher risk of developing coronary artery disease and myocardial infarction and up to an eight-fold higher risk of heart failure (3; 4). In a cohort-based analysis, a 1% increase in HbA1c corresponded to a 27% increase in cardiovascular event risk among patients with T2D (5). However, CVD risk following T2D diagnosis is heterogeneous and influenced by various risk factors in primary care setting (6; 7). For example, socio-demographic factors such as age (8), sex (9), marital status (10), and primary care location (11) have been linked to variation in risk of developing other chronic conditions. While individual risk factors and T2D-induced complications have been studied extensively, the probabilistic dependencies between these factors and post-diagnosis risk of CVD remain understudied. Furthermore, the blood glucose level –measured with a laboratory test called hemoglobin A1C (HbA1C) that captures the average blood glucose over the past three months (12) – at the time of T2D diagnosis is a critical quantitative indicator of early detection (13). Many individuals develop T2D years before it is clinically diagnosed (14). Delays in T2D detection often result in increased blood glucose (15; 16). Consequently, patients experiencing delays in T2D diagnosis often present with higher HbA1c levels at the time of diagnosis, indicating worsening organ damage (17). As routinely collected longitudinal electronic health records (EHR) data increasingly enable quantitative characterization of disease trajectories in primary care settings, HbA1c at diagnosis may serve as a clinically accessible surrogate for early disease detection and latent progression prior to diagnosis. However, the question remains whether the degree of HbA1c elevation at diagnosis – as a surrogate of early T2D detection – is linked to the subsequent risk of CVD in primary care settings.

Beyond early detection, early disease control is another key determinant of post-diagnosis outcomes. In the context of T2D, control refers the extent to which blood glucose levels are maintained within recommended clinical thresholds, commonly assessed using repeated HbA1c measurements. The first year after T2D diagnosis is increasingly recognized as a critical window during which effective T2D management can substantially alter the post-diagnosis health trajectories (18). Achieving and maintaining glycemic control during this period may reduce T2D-induced multi-organ system damage (19). Glycemic control is commonly characterized using glycemic exposure and stability. Glycemic exposure refers to the average level of blood glucose over time, reflecting the degree of chronic hyperglycemia experienced by the patient. In contrast, glycemic stability captures the extent to which blood glucose levels fluctuate across repeated measurements. Mean HbA1c provides a measure of average glycemic exposure over time (20), while variability in HbA1c, often quantified using the coefficient of variation (CV), captures fluctuations in control over time (21). Consistently high levels of blood glucose and greater variability in HbA1C have been associated with cardiovascular dysfunction (22; 23), suggesting that not only the level but also the consistency of control is important (23). As longitudinal EHR data increasingly support repeated assessment of disease progression after diagnosis, studying probabilistic dependencies between early detection (represented by the diagnostic lab value levels) and early post-diagnosis glycemic control to predict future CVD risk is a promising, yet underexplored, research direction.

Risk stratification in connection to the risk of developing CVD after T2D plays a central role in supporting early detection and post-diagnosis control of T2D. Stratifying patients according to their estimated risk enables clinicians to prioritize screening (24), tailor preventive strategies (25), and allocate resources to those most likely to benefit from early intervention (26). Because CVD risk evolves throughout the disease course, effective stratification frameworks ideally support assessment both at the time of diagnosis and during subsequent disease management. In primary care settings, routinely collected EHR data provide longitudinal demographic, behavioral, laboratory, and clinical information that can support repeated risk assessment over time. By combining these indicators into patient-specific risk estimates, stratification approaches support the identification of individuals who may otherwise remain undetected until more advanced stages of disease. These risk estimates are typically used to categorize patients into clinically meaningful groups, such as low-, intermediate-, or high-risk, based on thresholds informed by clinical knowledge (27; 28) or data-driven optimization methods (29; 30). However, developing modeling frameworks that can capture complex dependencies among multiple risk factors while supporting interpretable risk stratification for both early detection and post-diagnosis management in type 2 diabetes populations remains an ongoing challenge.

The objectives of this study are to quantify early detection and control of T2D using EHR data, and develop a temporally structured Bayesian Network (BN) to characterize the relationships among socio-demographics, early detection, early glycemic control, and post-diagnosis cardiovascular outcomes. Utilizing EHR data retrospectively derived from ten primary care locations over a time period of seven years, we develop, evaluate, and query a novel BN model capturing the dependencies among characteristics observed at different time points before and after the T2D diagnosis to enable patient-level cardiovascular risk stratification through probabilistic inference on the learned network.

## Related work

### Cardiovascular risk prediction in type 2 diabetes

A wide range of cardiovascular risk prediction models have been developed to estimate future cardiovascular events in individuals with and without T2D. Traditional statistical risk scores, such as the Systematic Coronary Risk Evaluation (SCORE2) (31), Framingham Risk Score (32), and Pooled Cohort Equations (33), are typically derived from large cohort studies and estimate 10-year cardiovascular risk using a limited set of clinical and non-clinical variables, including age, sex, systolic blood pressure, cholesterol levels, smoking status, and diabetes (34). Additionally, diabetes-specific risk engines, such as the UK Prospective Diabetes Study (UKPDS) risk model, have been proposed to better capture cardiovascular risk in individuals with T2D by incorporating disease-specific factors (35). More recently, machine learning approaches using EHR data have been introduced to improve predictive performance by leveraging larger sets of clinical variables and complex nonlinear relationships (36; 37). Despite these advances, several important limitations remain. Most traditional and machine learning models rely on baseline or aggregated predictors and treat risk factors as independent contributors to cardiovascular risk. As a result, they do not explicitly model the interdependencies among clinical variables, particularly during the early stages of disease. Moreover, early disease dynamics such as the interaction between timing of T2D detection and early glycemic control are rarely modeled jointly. These limitations hinder the ability of existing approaches to capture heterogeneous disease trajectories.

### Probabilistic graphical models in healthcare

Probabilistic graphical models, particularly BNs, have been widely used in healthcare to model uncertainty and complex relationships among clinical variables (38; 39; 40). BN represents a set of variables and their conditional dependencies through a directed acyclic graph (DAG), enabling probabilistic reasoning and inference (41). These models have been applied in a range of clinical settings, including decision support systems, diagnosis, and disease risk prediction (42; 43; 44). In clinical decision support, BNs facilitate reasoning over incomplete or uncertain data by integrating evidence from multiple sources and updating risk estimates as new information becomes available (45). They are particularly well-suited for jointly modeling risk factors, disease states, and outcomes (46). Despite these advantages, many applications of BNs in healthcare rely on expert knowledge or ad hoc model development processes, rather than leveraging EHR for structure learning (47).

Additionally, temporal extensions of BNs, such as dynamic Bayesian networks (DBNs), have been developed to model time-dependent processes by representing variables observed sequential and learning dependencies both within and between time points (48; 49). DBNs have been applied in healthcare to model disease progression, patient monitoring, and longitudinal clinical trajectories (50; 51). However, DBNs often require repeated observations at consistent time intervals, which commonly contradict clinical practice, particularly in primary care setting where patients may be observed in irregular time intervals.

### Bayesian networks for type 2 diabetes

BNs have been applied to T2D to model disease progression, complications, and metabolic risk (52; 53). Prior studies have used BNs to characterize relationships among glycemic measures, comorbidities, and the risk of diabetes-related complications, as well as to support clinical decision-making and risk prediction (54; 55). In addition, BN-based approaches have been explored for modeling broader cardiometabolic risk and estimating the likelihood of cardiovascular events in patients with T2D (56). However, existing studies typically focus on either baseline risk factors or subsequent complications, without jointly considering early disease processes. In particular, key determinants such as timing of T2D detection, early glycemic control, and demographic factors are rarely modeled together within a unified framework. Moreover, while temporal BN approaches have been proposed to model sequential clinical processes (50), few studies in T2D explicitly represent multiple disease stages within a single probabilistic framework. As a result, the joint probabilities of early detection, early glycemic control and subsequent cardiovascular risk represent a promising research domain to explore.

### Bayesian networks for clinical risk stratification

BN-based approaches have been applied to risk prediction and patient stratification across a range of conditions, including cardiovascular disease, pulmonary hypertension, oncology outcomes, and hospital-acquired infections (28; 29; 30; 57; 58). Despite the growing body of work on BN-based risk prediction, applications focusing specifically on CVD risk among individuals with T2D remain relatively limited. Much of the existing literature addresses broader populations or single-disease contexts, which may not fully capture the complex interactions among metabolic, clinical, and treatment-related factors that influence cardiovascular outcomes in T2D. Additionally, prior BN-based studies have not incorporated risk stratification across different stages of disease progression, such as at diagnosis, during the first year post-diagnosis, and over longer-term follow-up, thereby limiting their ability to support both early risk identification and post-diagnosis disease management.

## Materials and methods

### Data source and study population

This is an observational cohort study of adult patients diagnosed with T2D in the primary care setting. Diagnosis is represented by the first documentation of a relevant International Classification of Diseases version 10 (ICD-10) code in EHR during the study period (January 2017 - December 2023). S2 Appendix and S3 Appendix list all relevant ICD-10 codes used in this study. Data were derived from ten primary care provider (PCP) sites affiliated with a healthcare delivery system in the mid-Atlantic region of the United States and accessed between July 3, 2025 and June 6, 2026. The study cohort consisted of adults (age ≥ 18 years at index PCP visit) meeting the following criteria: ≥3 PCP visits during the study period, ≥1 T2D-related ICD-10 code, ≥3 post-diagnosis HbA1c laboratory results, and ≥1 year of follow-up observations after the T2D diagnosis to allow sufficient observations to quantify early detection and early control. Final sample size included 9,450 unique patients. The research protocol was approved by an Institutional Review Board (IRB), and the health system authorized the use of de-identified EHR data.

### Model development and validation

BN is defined as a pair *B* = (*G,* Θ), where *G* = (*V, E*) is a DAG consisting of nodes *V* = *{X*_1_, *X*_2_, … , *X_n_}* representing random variables and edges *E* representing direct probabilistic dependencies, and Θ is the set of conditional probability distributions (CPDs) associated with each node. The joint probability distribution over all variables factorizes according to the graph structure as

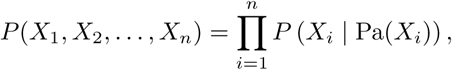

where Pa(*X_i_*) denotes the set of parent nodes of *X_i_* in *G*. The nodes *X_i_* correspond to the variables shown in Table 1.

**Table 1.** Nodes included in the OBBN and their corresponding layers.

| Node | Layer | Description | Unit / Category |
| --- | --- | --- | --- |
| Age | Layer 1 | Patient age at the time of T2D diagnosis | Quartiles (Q1–Q4) |
| Marital Status | Layer 1 | Marital status of the patient at baseline | Single / Married / Widowed |
| Sex | Layer 1 | Biological sex of the patient | Male / Female |
| PCP Location | Layer 1 | Primary care facility where the patient first received care | Categorical (facility location) |
| Prediabetes Diagnosis | Layer 2 | Indicator of whether the patient had a documented prediabetes diagnosis prior to T2D diagnosis | Binary (Yes/No) |
| HbA1c at Diagnosis | Layer 2 | Glycated hemoglobin level at the time of T2D diagnosis | Clinical categories (% HbA1c) |
| Delay Occurrence | Layer 2 | Indicator of delay in T2D diagnosis based on prior laboratory measurements | Binary (0/1) |
| Pre-T2D HbA1c Labs | Layer 2 | Number of HbA1c measurements before T2D diagnosis | Categories (0, 1, 2–3, 4+) |
| HgA1c CV (0-1 year) | Layer 3 | Coefficient of variation of HbA1c measurements during the first year after diagnosis, representing glycemic variability | Low / Moderate / High |
| mean HbA1c (0-1 year) | Layer 3 | Mean HbA1c level during the first year after diagnosis, representing average glycemic exposure | Clinical categories (% HbA1c) |
| CVD (0-1 year) | Layer 3 | Occurrence of cardiovascular disease within the first year after diagnosis | Binary (0/1) |
| HgA1c CV (1+ year) | Layer 4 | Coefficient of variation of HbA1c measurements after the first year of diagnosis | Low / Moderate / High |
| mean HbA1c (1+ year) | Layer 4 | Mean HbA1c level after the first year of diagnosis | Clinical categories (% HbA1c) |
| CVD (1+ year) | Layer 4 | Occurrence of cardiovascular disease after the first year following diagnosis | Categorical |

The variables listed in Table 1 were derived from EHR data using clinical guidance and prior literature (59; 60). To represent the temporal nature of the observation of these variables in clinical practice, variables were structured into four ordered *layers*, as listed below.

- Layer 1, referred to as baseline characteristics, represents information available prior to T2D diagnosis and includes demographic and care context variables: age at diagnosis, sex, marital status, and primary care facility. PCP location may indirectly capture geographic and socioeconomic factors such as neighborhood characteristics or access to healthcare (61).
- Layer 2 includes variables observed at or before T2D diagnosis, including HbA1c at diagnosis, documentation of prediabetes prior to T2D diagnosis, diagnostic delay occurrence, and the number of HbA1c measurements prior to diagnosis. Diagnostic delay was defined based on clinical input and ADA guidelines (62) using two criteria: (i) time from the first HbA1c in the diabetic range (≥ 6.5%) to diagnosis exceeding two months, or (ii) time between the first T2D-range HbA1c and the next HbA1c measurement exceeding six months.
- Layer 3 includes early post-diagnosis variables observed during the first year following diagnosis, including glycemic variability (HbA1c-CV), mean HbA1c, and occurrence of cardiovascular disease (CVD).
- Layer 4 presents the same variables observed beyond the first year after diagnosis: HbA1c-CV, mean HbA1c, and CVD occurrence. These variables are the outcomes of interest in this study.

The Order-Based Bayesian Network (OBBN) was specified as a discrete model and all variables were discretized and modeled using multinomial conditional probability distributions (CPDs) estimated from observed frequencies. Age at diagnosis was partitioned into quartiles based on the empirical distribution. Glycemic measures including HbA1c at diagnosis, mean HbA1c, and HbA1c-CV were discretized using clinically meaningful thresholds informed by prior studies (63; 64; 65) into the following categories: HbA1c at diagnosis (≤ 6.5%, 6.5–7%, 7–8%, and ¿8%), mean HbA1c (≤ 7%, 7.1–8%, and ¿8%), and HbA1c-CV (low, moderate, and high). The number of HbA1c measurements prior to diagnosis was discretized into ordinal categories (0, 1, 2–3, and ≥4), representing increasing levels of clinical monitoring. Missing laboratory-derived values were encoded as a separate “Not observed” category to preserve information on data availability.

Given the nodes, learning the structure (arcs) and parameters (conditional probabilities represented by the arcs) of a BN model is an NP-hard problem (66; 67). Structure learning can be categorized into constraint-based approaches which rely on conditional independence tests, score-based approaches which search over possible DAGs to optimize a scoring function, and hybrid approaches (68). In this study, the structure and parameter learning are achieved with purely data-driven structure learning using a hill-climbing search. This heuristic method iteratively adds, removes, or reverses arcs to optimize network fit to given data and identifies dependencies within the temporal ordering imposed by the layered framework. We chose this method for its computational efficiency and performance in learning BN structures from high-dimensional clinical data. Parameter learning was performed using the bnlearn package in R (RStudio version 2025.09.0 Build 387 (69)).

The data were partitioned into training, validation, and test sets using stratified sampling based on the long-term CVD outcome (CVD 1pyr) to preserve class proportions across splits, resulting in proportions of 60% training, 20% validation, and 20% test data. Model development was conducted on the training set. Internal model selection was performed using 5-fold cross-validation within the training data. In each fold, the BN structure was learned using hill-climbing search with a temporally constrained blacklist that prohibited arcs from later layers to earlier layers, thereby preserving the predefined temporal ordering of the OBBN framework and preventing information flow from future outcomes to earlier disease stages. Model parameters were estimated using Bayesian parameter learning. After cross-validation, the final OBBN was trained on the combined training and validation sets and then evaluated once on the held-out test set. This workflow was used to reduce optimistic bias and assess generalization to unseen patients.

### Performance evaluation

Model performance was assessed for the three Layer 4 outcomes (HbA1c CV, mean HbA1c, and CVD occurrence) representing disease progression after the first year post-diagnosis, up to 7 years of available study data. For each outcome node, posterior class probabilities were estimated at the individual patient level using likelihood-weighted sampling from the fitted OBBN. This approximate Bayesian inference approach generates samples conditional on observed evidence and weights them according to the likelihood of the observed data, enabling efficient estimation of posterior probabilities in complex networks with partially observed variables. Sampling was implemented using the cpdist function from the bnlearn package in R (RStudio version 2025.09.0 Build 387 (69)).

Predicted class labels were assigned according to the maximum a posteriori probability. Model performance was evaluated using log loss, multiclass Brier score, classification accuracy, and macro-averaged F1 score to assess both the probabilistic quality of predictions and classification performance. Log loss and multiclass Brier score were both used to evaluate probabilistic predictions because they capture complementary aspects of model calibration. Log loss assesses the extent to which predicted probabilities align with observed outcomes and is particularly sensitive to highly confident incorrect predictions, whereas the Brier score measures the overall squared difference between predicted probabilities and observed outcomes across all classes (70; 71). For both metrics, lower values indicate better calibrated and more reliable probabilistic predictions, with values approaching zero reflecting optimal performance. Classification accuracy quantified the proportion of correctly classified observations among all predictions, with higher values indicating stronger overall predictive performance (72). Because accuracy may be influenced by class imbalance, the macro-averaged F1 score was additionally used to provide a more balanced assessment across outcome classes. Macro-F1 summarizes the harmonic mean of precision and recall calculated separately for each class and then averaged equally across classes, with values closer to 1 indicating better discrimination across all outcome categories, including less frequent classes (73). Together, these metrics provide complementary evaluation of the OBBN model by assessing calibration, probabilistic reliability, and classification performance across multiclass outcomes.

For comparative evaluation, multinomial logistic regression and random forest models were trained separately for each outcome using the same predictors and data splits. Predicted probabilities from these models were evaluated using the same metrics to enable direct comparison with the OBBN approach.

### Two-phase risk stratification

Risk estimation was conducted in two sequential phases reflecting early (Phase 1, referring to 0-1 year post T2D diagnosis) and long-term (Phase 2, referring to *>* 1 year post T2D diagnosis) prediction windows. For each outcome, predicted risk was defined as the posterior probability of the clinically adverse category of outcome nodes. To characterize variation in predicted risk, patients were stratified according to the distribution of predicted probabilities. Percentile-based thresholds were used to define risk strata: individuals with predicted probabilities in the bottom decile (≤ 10*^th^* percentile) were classified as low risk, those in the top decile (≥ 90*^th^* percentile) were classified as high risk, and the remaining individuals were considered to have intermediate risk. These thresholds were applied consistently across all outcomes and across both phases of risk estimation.

### Phase 1: Early glycemic control and early cardiovascular risk

In Phase 1, we estimated the probability of outcomes occurring during the first year following T2D diagnosis. These outcomes included indicators of glycemic control (mean HbA1c and HbA1c CV) and CVD events observed during the first year after T2D diagnosis. Patient-specific posterior probabilities were estimated using Layer 1 and Layer 2 nodes that described demographic characteristics and information available at the time of diagnosis. The OBBN was queried to estimate the likelihood of adverse early glycemic control and development of CVD. Posterior probabilities were estimated for the categories of each outcome (HbA1c ≥ 8%, high HbA1c coefficient of variation, and presence of CVD). Risk stratification was applied to each outcome using the percentile-based thresholds to capture differences between low-risk and high-risk populations.

### Phase 2: Estimation of long-term glycemic control and cardiovascular risk

In Phase 2, we estimated longer-term clinical risks occurring after the first year following T2D diagnosis. In addition to baseline characteristics, the model incorporated early clinical outcomes observed during the first year. Specifically, the Layer 3 nodes representing early outcomes (mean HbA1c category, HbA1c CV category, and CVD within the first year) were included in the inference process. This approach allowed quantifying the relationship between early indicators of glycemic control and CVD events and later outcomes, thereby reflecting the temporal dependencies inherent to T2D disease progression. Posterior probabilities were estimated for the corresponding long-term outcomes (mean HbA1c ≥ 8%, high HbA1c CV, and CVD occurring after the first year), and patients were stratified using the same percentile-based thresholds as described above.

### Comparison between early and later predicted risks

Statistical analyses were conducted to evaluate differences in predicted risk distributions across the two phases of the model. To assess differences between early and later predicted risks, predicted probabilities of Layer 3 and Layer 4 nodes were compared within the same individuals for each outcome. Because predictions were generated for the same individuals, the Wilcoxon signed-rank test, a nonparametric test for paired samples, was used to account for within-subject correlation and to assess differences in median predicted probabilities across phases. Differences between high-risk and low-risk groups were evaluated using the Wilcoxon rank-sum test (Mann–Whitney U test), which compares the distributions of two independent groups. In addition, the Kolmogorov–Smirnov (KS) test was used to compare the overall distributions of predicted probabilities between groups. Two-sided statistical tests were used, and statistical significance was defined as *p <* 0.05.

## Results

### Study population characteristics

Table 2 summarizes the cohort characteristics. The study population was predominantly female (61.9%), with most individuals being single or married. A relatively small proportion (14.6%) had documented prediabetes prior to T2D diagnosis, while 42.0% experienced delay occurrence. Notably, HbA1c at diagnosis was frequently missing (45.7%), though among observed values, a substantial fraction (17.1%) presented with poor glycemic control (*>*8.0%). Early post-diagnosis outcomes (0–1 year) indicate that approximately half of patients achieved target glycemic control (HbA1c ≤ 7%), yet a considerable proportion (23.6%) remained poorly controlled. HbA1c variability was predominantly low among observed cases, though missingness remained substantial. In contrast, long-term outcomes (¿1 year) demonstrate a shift toward increased glycemic variability and a higher burden of cardiovascular disease, suggesting worsening glycemic stability over time despite modest improvements in mean HbA1c control.

**Table 2.** Study population characteristics. Categorical variables are reported as n (%). PCP stands for primary care provider, T2D for Type 2 Diabetes, CVD for Cardiovascular Disease and CV for coefficient of variation.

| Characteristic | N (%) |
| --- | --- |
| **Layer 1: Demographics** |  |
| **Sex** |  |
| Female | 5854 (61.9%) |
| Male | 3596 (38.1%) |
| **Marital Status** |  |
| Married | 3607 (38.2%) |
| Single | 4557 (48.2%) |
| Widowed | 1286 (13.6%) |
| **Layer 2: Diagnosis Stage** |  |
| **HbA1c at Diagnosis (%)** |  |
| ≤ 6.5 | 1377 (14.6%) |
| 6.5 – 7.0 | 1063 (11.3%) |
| 7.0 – 8.0 | 1070 (11.3%) |
| > 8.0 | 1619 (17.1%) |
| Not observed | 4321 (45.7%) |
| **Delay Occurrence** |  |
| No Delay | 5483 (58.0%) |
| Delay | 3967 (42.0%) |
| **Layer 3: Early Post-Diagnosis Outcomes (0–1 year)** |  |
| **HbA1c CV** |  |
| Low | 3422 (36.2%) |
| Moderate | 1162 (12.3%) |
| High | 929 (9.8%) |
| Not Observed | 3937 (41.7%) |
| **Mean HbA1c (%)** |  |
| ≤ 7 | 4650 (49.2%) |
| 7.1 – 8 | 1652 (17.5%) |
| > 8 | 2229 (23.6%) |
| Not Observed | 919 (9.7%) |
| **CVD** |  |
| 0 | 7800 (82.5%) |
| 1 | 1650 (17.5%) |
| **Layer 4: Long-term Post-Diagnosis Outcomes (1+ year)** |  |
| **HbA1c CV** |  |
| Low | 3287 (34.8%) |
| Moderate | 2561 (27.1%) |
| High | 2987 (31.6%) |
| Not Observed | 615 (6.5%) |
| **Mean HbA1c (%)** |  |
| ≤ 7 | 4781 (50.6%) |
| 7.1 – 8 | 2139 (22.6%) |
| > 8 | 2470 (26.1%) |
| Not Observed | 60 (0.7%) |
| **CVD** |  |
| 0 | 6083 (64.4%) |
| 1 | 1717 (18.2%) |
| 2 | 1650 (17.4%) |
| **Total number of unique patients** | **9450 (100%)** |

### Learned network structure

The final network contained 14 nodes and 20 directed arcs spanning demographic characteristics, diagnostic-stage factors, early glycemic and cardiovascular outcomes, and long-term outcomes as shown in Figure 1.

**Fig 1.**
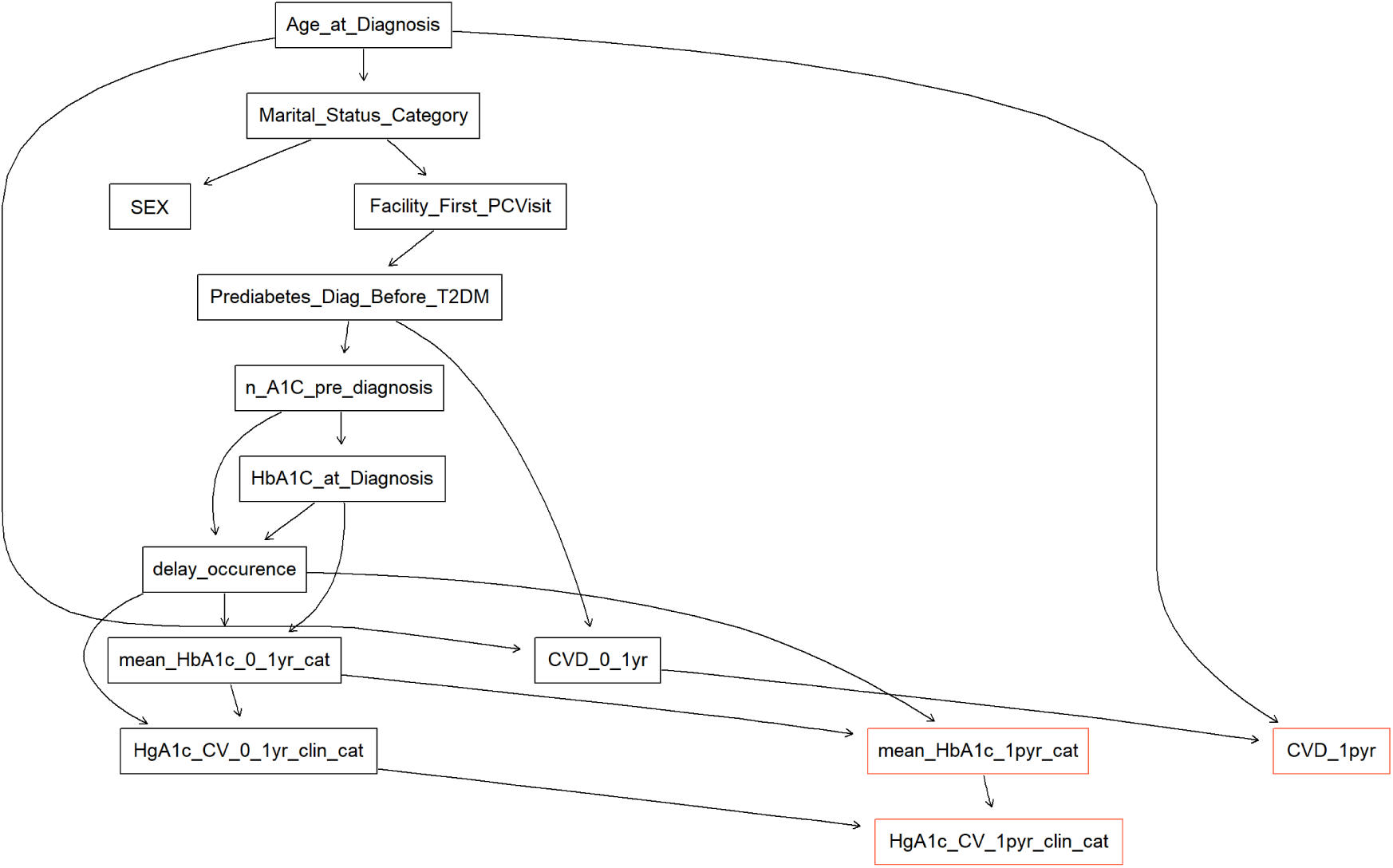
Final structure of the learned Order-Based Bayesian Network (OBBN) where the arcs and parameters are learned from data. The network comprises 14 nodes and 20 arcs. Nodes correspond to the variables listed in Table 1. Red outlined nodes represent the outcomes of the study.

### Predictive performance

Table 3 describes the comparative performance of the OBBN, multinomial logistic regression, and random forest models across all Layer 4 nodes as outcomes. For long-term CVD occurrence, OBBN achieved the highest accuracy (0.818) and macro F1 score (0.938), indicating strong discrimination across outcome classes. Multinomial logistic regression yielded slightly lower log loss (0.406) and Brier score (0.265), suggesting marginally improved probabilistic calibration. For both HbA1c-related outcomes, i.e. HbA1c variability (¿1 year) and mean HbA1c (¿1 year), performance differences across models were modest, with OBBN presenting results comparable to alternative approaches across evaluation metrics.

**Table 3.** Comparison of model performance across outcomes.

| Model | Node | Log Loss | Brier Score | Accuracy | Macro F1 |
| --- | --- | --- | --- | --- | --- |
| OBBN | CVD (1+ year) | 0.426 | 0.277 | 0.818 | 0.938 |
| Random Forest | CVD (1+ year) | 0.434 | 0.274 | 0.814 | 0.661 |
| Multinomial Logistic | CVD (1+ year) | 0.406 | 0.265 | 0.812 | 0.651 |
| OBBN | HbA1c CV (1+ year) | 1.002 | 0.558 | 0.587 | 0.446 |
| Random Forest | HbA1c CV (1+ year) | 0.971 | 0.538 | 0.593 | 0.479 |
| Multinomial Logistic | HbA1c CV (1+ year) | 0.947 | 0.526 | 0.602 | 0.494 |
| OBBN | mean HbA1c (1+ year) | 0.846 | 0.482 | 0.643 | 0.580 |
| Random Forest | mean HbA1c (1+ year) | 0.716 | 0.415 | 0.679 | 0.587 |
| Multinomial Logistic | mean HbA1c (1+ year) | 0.710 | 0.413 | 0.685 | 0.597 |

### Two phase risk stratification

#### Phase 1 results

Conditional BN queries using selected Layer 2 early detection indicators demonstrated substantial differences in posterior probabilities for several first-year outcomes. Patients with HbA1c at diagnosis *>* 8% had higher predicted probabilities of first-year mean HbA1c *>* 8% compared with those with HbA1c ≤ 6.5% at diagnosis (53.3% vs. 19%). Similarly, patients with HbA1c at diagnosis *>* 8% had higher predicted probabilities of first-year high HbA1c CV than those with HbA1c ≤ 6.5% (18.7% vs. 3.1%). In contrast, posterior probabilities for first-year CVD occurrence differed only modestly according to HbA1c at diagnosis (17.6% vs. 15.9%). Delay occurrence showed comparatively smaller differences in predicted probabilities across outcomes, including first-year mean HbA1c *>* 8% (24.1% vs. 22.8%), high HbA1c CV (6.2% vs. 12.6%), and CVD occurrence (17% vs. 18%).

Predicted probability distributions for early clinical outcomes during the first year following diagnosis are shown in Figure 2. Kernel density estimates demonstrate clear separation between predefined high-risk and low-risk groups across all outcomes. For CVD within 1 year post T2D diagnosis, the high-risk group (N=193) had a mean predicted probability of 28.9% (median 28.8%, SD=0.006), whereas the low-risk group (N=192) had substantially lower predicted probabilities (mean 5.2%, median 6.5%, SD=0.024). Individuals in the middle-risk group (N=1506) showed intermediate predicted probabilities (mean 17.5%, median 17.7%, SD=0.062). For high HbA1c CV, the high-risk group (N=191) had a mean predicted probability of 21.7% (median 22.3%, SD=0.025), compared with 2.5% in the low-risk group (N=193; median 2.5%, SD=0.003). The middle-risk group (N=1507) again showed intermediate values (mean 8.9%, median 8.6%, SD=0.033). For mean HbA1c ≥ 8%, predicted probabilities were substantially higher in the high-risk group (mean 55.1%, median 55.6%, SD=0.025) than in the low-risk group (mean 1.6%, median 1.7%, SD=0.002), with intermediate probabilities observed in the middle-risk group (mean 22.2%, median 24.3%, SD=0.115).

**Fig 2.**
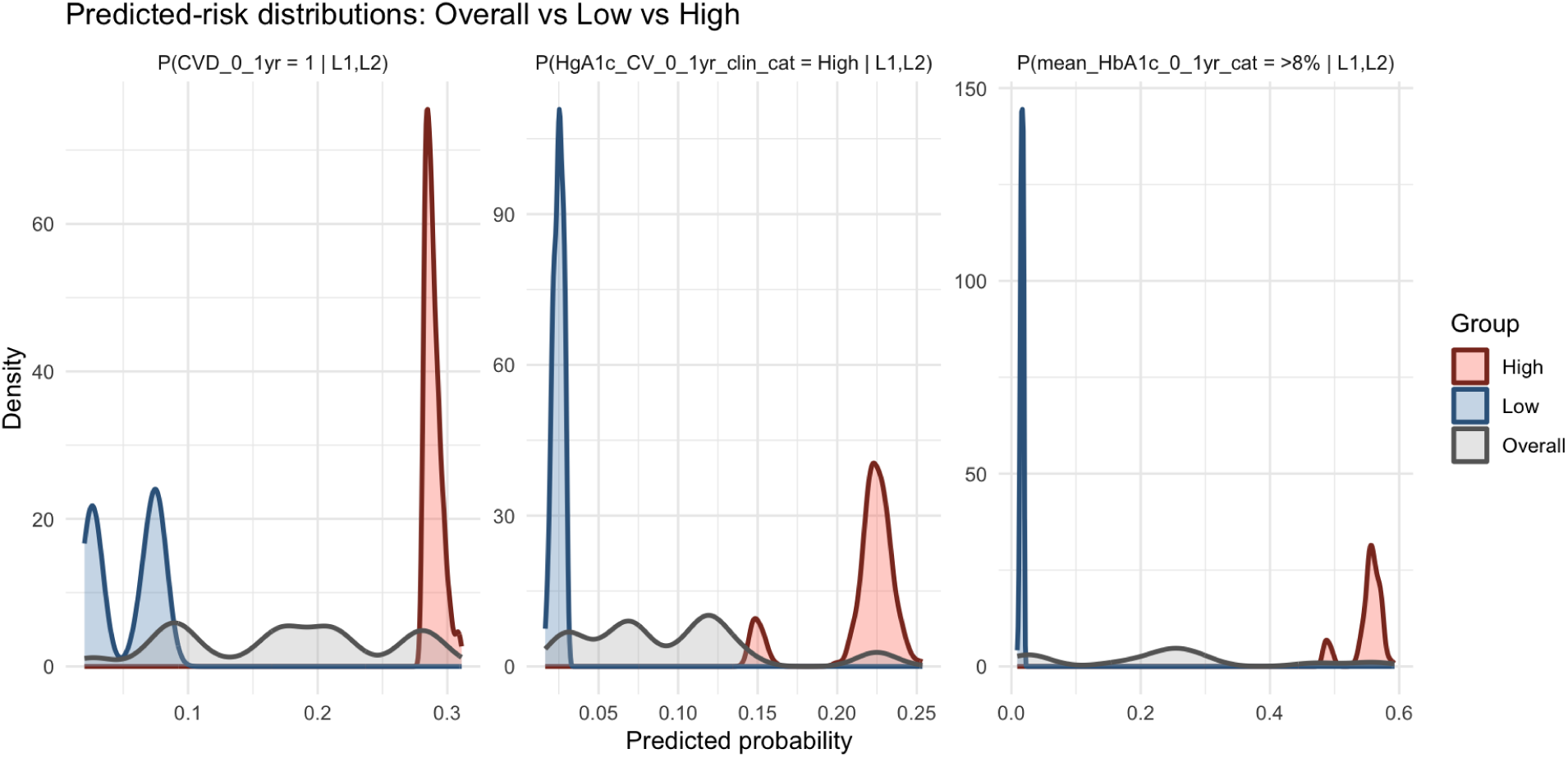
Predicted probabilities for early outcomes during the first year after diagnosis. Kernel density plots show the distribution of patient-level predicted probabilities for early CVD events, high HbA1c CV, and mean HbA1c ≥ 8%, estimated using baseline demographic and diagnostic information (Layers 1–2). Curves represent the overall cohort (gray) and high-risk (red) and low-risk (blue) groups defined by the top and bottom deciles of predicted risk.

### Phase 2 results

Predicted probability distributions for long-term outcomes incorporating early clinical evidence are shown in Figure 3. Similar separation between high- and low-risk groups was observed. For long-term CVD, predicted probabilities were near zero in the low-risk group (N=330; mean 0.0, median 0.0), whereas the high-risk group (N=191) had a mean predicted probability of 33.5% (median 33.4%). For long-term HbA1c CV, predicted probabilities averaged 11.8% in the low-risk group (N=194; median 12.0%) and 56.8% in the high-risk group (N=190; median 56.8%). Similarly, for long-term mean HbA1c ≥ 8%, predicted probabilities were 4.3% in the low-risk group (N=207; median 4.4%) and 0.647 in the high-risk group (N=195; median 64.6%).

**Fig 3.**
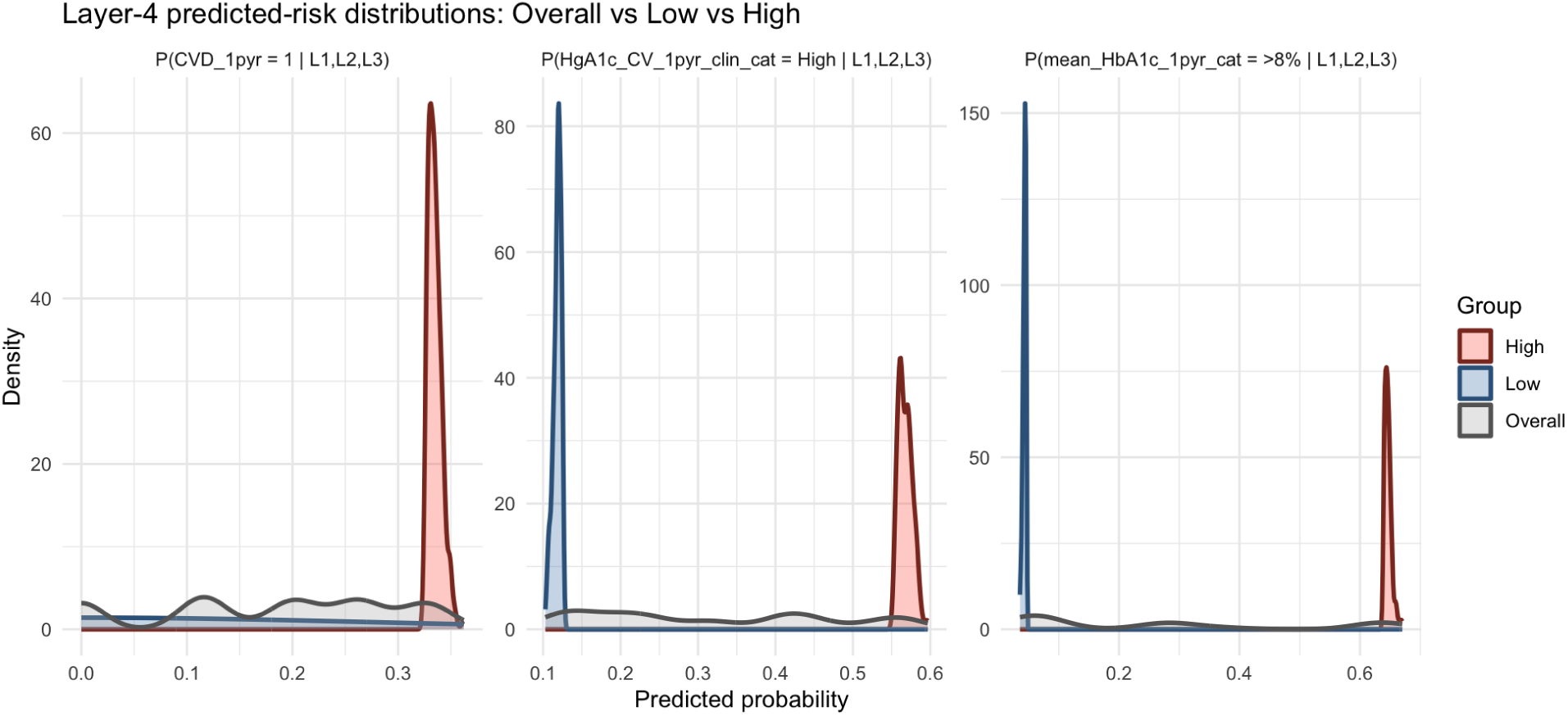
Predicted probabilities for long-term outcomes after the first year following diagnosis. Kernel density plots show the distribution of predicted probabilities for long-term CVD events, high HbA1c CV, and mean HbA1c ≥ 8% occurring after the first year. Predictions incorporate baseline characteristics and early clinical outcomes (Layers 1–3). Curves represent the overall cohort (gray) and high-risk (red) and low-risk (blue) groups defined by the top and bottom deciles of predicted risk.

### Comparison between early and later predicted risks of CVD

Figure 4 is a boxplot that shows the distribution of predicted probabilities across model layers for each outcome. Predicted risks differed significantly between early (Layer 3) and later (Layer 4) estimates across all outcomes, with Layer 4 generally showing higher median risk estimates and greater variability. For HbA1c CV, the median predicted risk increased from approximately 0.08 in Layer 3 to 0.30 in Layer 4, representing an approximate 275% increase, alongside a substantial widening of the interquartile range. For CVD occurrence, the median predicted risk increased modestly from approximately 0.18 to 0.20 (approximately 11%), with slightly greater spread in Layer 4. Similarly, for mean HbA1c, the median predicted risk increased from approximately 0.24 to 0.25, while the interquartile range expanded from approximately 0.04–0.29 to 0.04–0.38.

**Fig 4.**
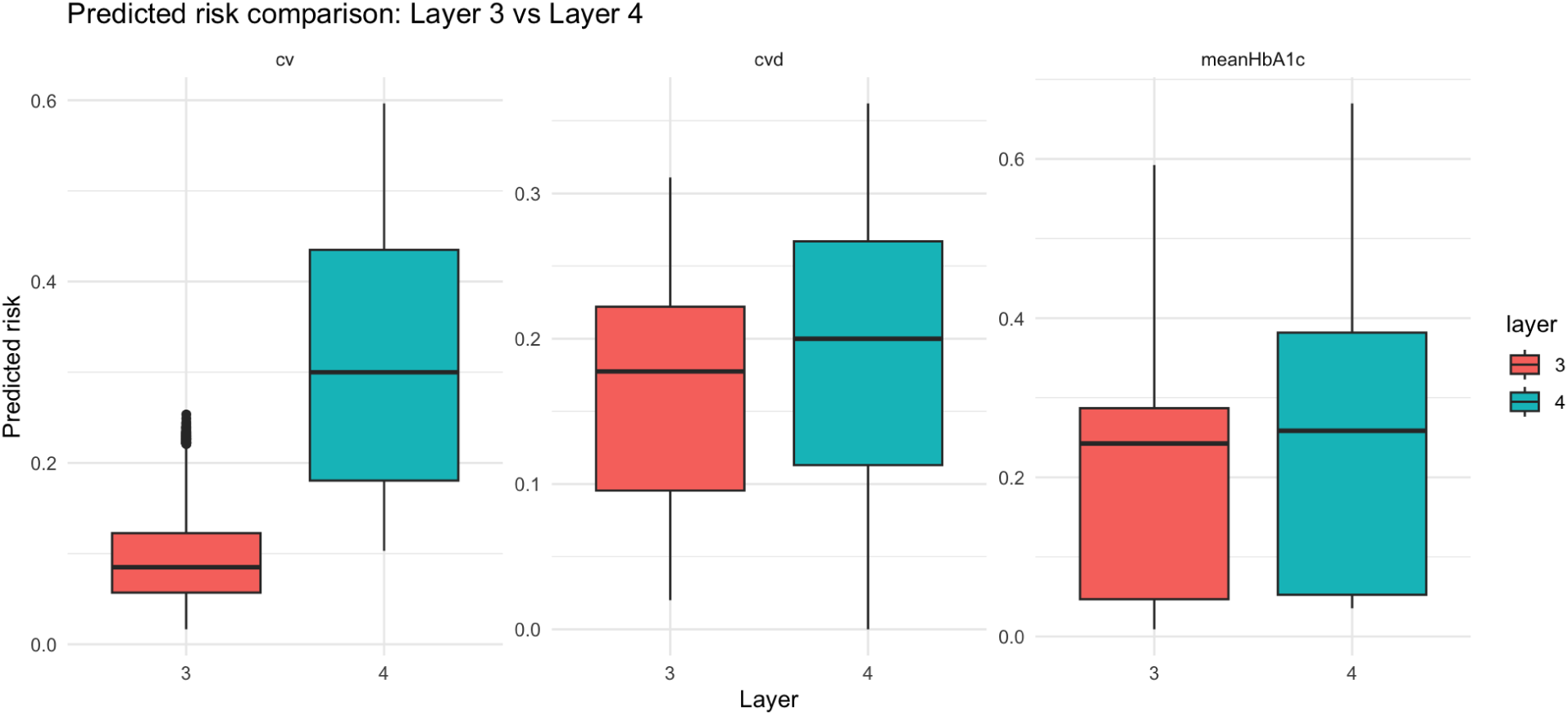
Comparison of predicted risks across model layers. Boxplots show patient-level predicted probabilities for CVD, HbA1c CV, and mean HbA1c ≥ 8% estimated using Layer 3 and Layer 4 of the model, illustrating the distribution of predicted risks for each outcome across layers.

Wilcoxon signed-rank tests indicated significant differences for mean HbA1c (*p <* 0.001), HbA1c CV (*p <* 0.001), and CVD risk (*p <* 0.001). In addition, comparisons between low-risk and high-risk groups for long-term outcomes showed significant differences in predicted risk distributions based on both Wilcoxon rank-sum and Kolmogorov–Smirnov tests (all p*<* 0.001).

## Discussion

### Clinical implications

This study aimed to characterize the relationships between socio-demographic, early detection, early glycemic control, and post-diagnosis outcomes using a temporally structured BN. The results have several clinical implications for patients diagnosed with T2D in primary care setting.

First, the model demonstrated substantial heterogeneity in predicted risk for both glycemic control outcomes and CVD following diagnosis. For example, predicted probabilities of early CVD ranged from a mean of 5.2% in the low-risk group to 28.9% in the high-risk group, while predicted probabilities of mean HbA1c ≥ 8% during the first year ranged from 1.6% in the low-risk group to 55.1% in the high-risk group. Similar separation was observed for HbA1c variability (2.5% vs. 21.7%). These differences indicate that baseline characteristics at the time of diagnosis can differentiate patients regarding their expected clinical trajectories. Early identification of individuals at elevated risk may support targeted monitoring and proactive management strategies, consistent with guideline recommendations emphasizing individualized cardiovascular and metabolic risk assessment in patients with T2D. Specifically, the proposed model could support clinicians in making risk-informed referral and treatment decisions by identifying patients at higher predicted risk of poor glycemic control or CVD who may benefit from earlier endocrinology or cardiology referral, more frequent monitoring, medication optimization, and closer follow-up.

Second, incorporating early clinical outcomes into the model changed predicted risks for later outcomes. When early indicators of glycemic control and CVD were included, predicted long-term risks showed significant differences between risk groups. For example, predicted probabilities of later CVD were near zero in the low-risk group, but reached approximately 0.335 in the high-risk group. Similarly, predicted risks for long-term HbA1c CV increased from a mean of 0.118 in the low-risk group to 0.568 in the high-risk group, while predicted risks for mean HbA1c ≥ 8% increased from 0.043 to 0.647 across the same groups. These findings support previous evidence linking early glycemic patterns and variability with later complications in T2D. From a clinical perspective, the proposed two-phased model could enable clinicians to estimate risk at the time of T2D diagnosis and subsequently update these assessments after the first year post-diagnosis as clinical outcomes become available over the course of disease progression.

Third, the operational definition of “delay” developed in this study provides a potential indicator of barriers to timely care. Delays between abnormal HbA1c measurements, diagnosis confirmation, and subsequent clinical management may contribute to poorer metabolic control. By incorporating delay as a surrogate derived from data, the model identifies patients who may have experienced gaps in early care processes. Such indicators could help clinicians recognize individuals who require closer follow-up or earlier intervention.

Finally, the findings highlight the potential value of probabilistic graphical models as interpretable decision-support tools for CVD risk assessment among patients with T2D in primary care. The OBBN framework enables estimation of patient-specific probabilities of cardiovascular events while explicitly representing dependencies between baseline characteristics, early glycemic control, and subsequent cardiovascular outcomes. Bayesian network–based approaches have previously been proposed as interpretable tools for clinical decision support in complex chronic diseases (54; 55; 56). In the context of T2D care, such models may complement existing CVD risk assessment strategies by providing probabilistic, patient-specific estimates that incorporate both baseline characteristics and early post-diagnosis disease trajectories.

### Methodological contributions

The novelty of the developed OBBN lies in the four-layer structure and its ability to capture the probabilistic dependencies between critical, yet understudied, delay, control and CVD trajectories. The order-based construction constrained parent–child relationships to a clinically plausible sequence. This predefined the network structure ensured that the final model more closely reflected real-world diagnostic processes. Rather than producing only patient-level predictions, the model supports conditional probability queries that estimate outcome risk under specified combinations of clinical states. This capability allows investigators to explore how alternative early disease trajectories such as delayed detection or suboptimal post-diagnosis glycemic control may influence long-term outcomes.

Importantly, the OBBN encodes clinically meaningful temporal and probabilistic relationships to support temporal risk updating across disease stages. Although traditional models showed slight advantages in some metrics, they lack this structured interpretability. By structuring the network into sequential layers corresponding to baseline characteristics, early clinical indicators, and later outcomes, the model allows risk estimates to evolve as new clinical evidence becomes available. This approach reflects the progressive nature of T2D and CVD risk development and provides a framework for integrating early disease signals into later risk prediction. Such dynamic probabilistic modeling may offer advantages over static prediction models by enabling both patient-level risk estimation and population-level stratification based on evolving clinical trajectories.

Additionally, the OBBN demonstrated predictive performance comparable to multinomial logistic regression and random forest across all outcomes, with particularly strong discrimination for CVD, while differences in calibration were modest. The overall moderate performance across models suggests that additional unmeasured factors may contribute to outcome variability, highlighting the need for incorporating broader clinical, behavioral, and contextual variables in future work.

### Study limitations

Several limitations should be considered when interpreting the findings of this study. First, the analysis was restricted to data from ten PCP sites within the same United States healthcare delivery system, which may limit the generalizability of the results to other populations or care settings.

Second, diagnostic delay was operationalized using a surrogate definition based on EHR data, as the exact onset of hyperglycemia and date of first elevated HbA1c were not directly recorded; this approximation may differ from the true delay experienced by patients. Additionally, definition of “delay” related to T2D diagnosis required iterative refinement through close collaboration with clinicians. For each patient, the first relevant ICD code (documented during the study period) represented the time of formal diagnosis, and this time point was compared to the first documentation of elevated HbA1c time point to determine the occurrence of delay. A critical limitation of this approach to operationalize diagnostic delay is that both “first elevated HbA1c” and “first ICD code” rely on manual transcription of certain measurements and input of a formal diagnosis code in data by clinicians. Manual data entry in EHR systems is prone to errors and delays (74). Therefore, we reviewed the identified delay occurrences with two primary care clinicians (W.G., E.S.) during regular meetings. In 22 cases, the data were further evaluated through iterative chart review to confirm the occurrence of delay. Other data limitations which required clinical input included infeasible or inconsistent lab values in case of multiple measurements on the same date as well as transcribed lab values measured prior to the visit of data entry. This emphasizes the critical role of clinician collaborators for their input with variable definition, the importance of transparent preprocessing workflows, and the need for reproducible documentation of data sources and processing steps in multimodal health data research.

Third, causal claims cannot be made regarding relationships among variables in the BN. The OBBN model represents probabilistic dependencies and conditional associations learned from clinician knowledge and observational data, rather than cause-effect relationships. The retrospective, non-interventional nature of the data further limits causal interpretation due to potential confounding and unobserved variables.

Accordingly, the models are intended for probabilistic reasoning under uncertainty, not causal discovery. Moreover, there is an absence of medication and treatment data and without information on treatment intensity, frequency, and adherence, it is difficult to fully interpret measures of disease control derived from clinical and laboratory observations.

### Recommendations for future research

Future research can expand this work in several directions. First, efforts are needed to extend the study population to larger and more diverse cohorts, incorporating additional social and behavioral determinants of health. Furthermore, quantifying the economic implications of delayed diagnosis would provide a foundation for evaluating targeted interventions. This is particularly important in value-based care environments, where financial sustainability must be balanced with clinical outcomes. Another direction for future work could focus on the impact of multi-morbidity. Many individuals with T2D either present with or eventually develop other chronic conditions in addition to CVD, such as chronic kidney disease, or hypertension. These comorbidities can influence patterns of diagnostic delay, alter trajectories of glycemic control and CVD, and complicate care delivery in ways not addressed in the current study. Incorporating multimorbidity into future models would provide a more comprehensive view of patient trajectories and better reflect the clinical complexity of real-world diabetes populations. Future studies should integrate clinical diagnostic data and laboratory observations with treatment and medication data to provide a more comprehensive understanding of disease control. Finally, there is an opportunity to investigate the transferability of this modeling framework to other chronic diseases prone to diagnostic delay and long-term disease monitoring.

## Conclusion

This study presents an OBBN framework for modeling probabilistic dependencies across early detection, early glycemic control, and CVD risk following T2D diagnosis. By organizing variables into sequential layers that reflect clinically plausible disease trajectories, the model captures temporal relationships among demographic characteristics, diagnostic-stage factors, early disease management, and subsequent outcomes. The framework enables estimation of patient-specific probabilities of adverse outcomes and supports risk stratification based on predicted probabilities. Applied to a real-world primary care cohort, the OBBN achieved predictive performance comparable to conventional machine learning approaches while providing interpretable probabilistic representations of disease progression. The model revealed substantial heterogeneity in predicted risks for both glycemic control outcomes and CVD and demonstrated clear separation between high-risk and low-risk groups. By incorporating early clinical indicators into subsequent predictions, the framework allows risk estimates to evolve as additional evidence about early disease trajectories becomes available. Overall, these findings highlight the potential of structured probabilistic graphical models to support interpretable cardiovascular risk stratification among patients with T2D in primary care settings. The proposed OBBN framework complements existing cardiovascular risk assessment approaches by probabilistically modeling how baseline characteristics and early glycemic trajectories influence subsequent CVD risk. Future work should incorporate additional clinical, behavioral, and contextual variables to further improve predictive performance and expand the applicability of probabilistic modeling frameworks for longitudinal risk assessment and clinical decision support.

## Data Availability

The minimal dataset and accompanying code are publicly available on Synapse (Sage Bionetworks) at https://www.synapse.org/Synapse:syn75373876/wiki/641088.

## Acknowledgments

The authors would like to thank the clinicians at MedStar Georgetown University Hospital and data scientists at MedStar Health Research Institute, particularly Laura Schubel for project management support and Sonita S. Bennett for data acquisition and cleaning. This project was funded under grant number 1R01HS029792-01 from the Agency for Healthcare Research and Quality (AHRQ), U.S. Department of Health and Human Services (HHS). The authors are solely responsible for this document’s contents, findings, and conclusions, which do not necessarily represent the views of AHRQ or HHS.

## Supporting information

### S1 Appendix. Table of Acronyms

| Acronym | Definition |
| --- | --- |
| ADA | American Diabetes Association |
| BN | Bayesian Network |
| CV | Coefficient of Variation |
| EN | Elastic Net regression |
| HbA1c | Hemoglobin A1c |
| ICD-10 | International Classification of Diseases, Tenth Revision |
| OBBN | Order-Based Bayesian Network |
| PCP | Primary Care Provider |
| $R^2$ | Coefficient of Determination |
| RF | Random Forest |
| RMSE | Root Mean Squared Error |
| T2D | Type 2 Diabetes |

### S2 Appendix. List of ICD-10 Codes for Type 2 Diabetes Diagnosis

| Number | ICD Code | Description |
| --- | --- | --- |
| 1 | E11.00 | Type 2 diabetes mellitus with hyperosmolarity without nonketotic hyperglycemic-hyperosmolar coma (NKHHC) |
| 2 | E11.01 | Type 2 diabetes mellitus with hyperosmolarity with coma |
| 3 | E11.10 | Type 2 diabetes mellitus with ketoacidosis without coma |
| 4 | E11.21 | Type 2 diabetes mellitus with diabetic nephropathy |
| 5 | E11.22 | Type 2 diabetes mellitus with diabetic chronic kidney disease |
| 6 | E11.29 | Type 2 diabetes mellitus with other diabetic kidney complication |
| 7 | E11.319 | Type 2 diabetes mellitus with unspecified diabetic retinopathy with macular edema |
| 8 | E11.3211 | Type 2 diabetes mellitus with mild nonproliferative diabetic retinopathy with macular edema, right eye |
| 9 | E11.3212 | Type 2 diabetes mellitus with mild nonproliferative diabetic retinopathy with macular edema, left eye |
| 10 | E11.3213 | Type 2 diabetes mellitus with mild nonproliferative diabetic retinopathy with macular edema, bilateral |
| 11 | E11.3219 | Type 2 diabetes mellitus with mild nonproliferative diabetic retinopathy with macular edema, unspecified eye |
| 12 | E11.3291 | Type 2 diabetes mellitus with mild nonproliferative diabetic retinopathy without macular edema, right eye |
| 13 | E11.3292 | Type 2 diabetes mellitus with mild nonproliferative diabetic retinopathy without macular edema, left eye |
| 14 | E11.3293 | Type 2 diabetes mellitus with mild nonproliferative diabetic retinopathy without macular edema, bilateral |
| 15 | E11.3299 | Type 2 diabetes mellitus with mild nonproliferative diabetic retinopathy without macular edema, unspecified eye |
| 16 | E11.3311 | Type 2 diabetes mellitus with moderate nonproliferative diabetic retinopathy with macular edema, right eye |
| 17 | E11.3313 | Type 2 diabetes mellitus with moderate nonproliferative diabetic retinopathy with macular edema, bilateral |
| 18 | E11.3391 | Type 2 diabetes mellitus with moderate nonproliferative diabetic retinopathy without macular edema, right eye |
| 19 | E11.3392 | Type 2 diabetes mellitus with moderate nonproliferative diabetic retinopathy without macular edema, left eye |
| 20 | E11.3393 | Type 2 diabetes mellitus with moderate nonproliferative diabetic retinopathy without macular edema, bilateral |
| 21 | E11.3399 | Type 2 diabetes mellitus with moderate nonproliferative diabetic retinopathy without macular edema, unspecified eye |
| 22 | E11.3411 | Type 2 diabetes mellitus with severe nonproliferative diabetic retinopathy with macular edema, right eye |
| 23 | E11.3412 | Type 2 diabetes mellitus with severe nonproliferative diabetic retinopathy with macular edema, left eye |
| 24 | E11.3413 | Type 2 diabetes mellitus with severe nonproliferative diabetic retinopathy with macular edema, bilateral |
| 25 | E11.3419 | Type 2 diabetes mellitus with severe nonproliferative diabetic retinopathy with macular edema, unspecified eye |
| 26 | E11.3492 | Type 2 diabetes mellitus with severe nonproliferative diabetic retinopathy without macular edema, left eye |
| 27 | E11.3493 | Type 2 diabetes mellitus with severe nonproliferative diabetic retinopathy without macular edema, bilateral |
| 28 | E11.3499 | Type 2 diabetes mellitus with severe nonproliferative diabetic retinopathy without macular edema, unspecified eye |
| 29 | E11.3511 | Type 2 diabetes mellitus with proliferative diabetic retinopathy with macular edema, right eye |
| 30 | E11.3513 | Type 2 diabetes mellitus with proliferative diabetic retinopathy with macular edema, bilateral |
| 31 | E11.3519 | Type 2 diabetes mellitus with proliferative diabetic retinopathy with macular edema, unspecified eye |
| 32 | E11.3521 | Type 2 diabetes mellitus with proliferative diabetic retinopathy without macular edema, right eye |
| 33 | E11.3523 | Type 2 diabetes mellitus with proliferative diabetic retinopathy without macular edema, bilateral |
| 34 | E11.3532 | Type 2 diabetes mellitus with diabetic macular edema, resolved following treatment, left eye |
| 35 | E11.3533 | Type 2 diabetes mellitus with diabetic macular edema, resolved following treatment, bilateral |
| 36 | E11.3542 | Type 2 diabetes mellitus with diabetic cataract, left eye |
| 37 | E11.3551 | Type 2 diabetes mellitus with stable proliferative diabetic retinopathy without macular edema, right eye |
| 38 | E11.3552 | Type 2 diabetes mellitus with stable proliferative diabetic retinopathy without macular edema, left eye |
| 39 | E11.3553 | Type 2 diabetes mellitus with stable proliferative diabetic retinopathy without macular edema, bilateral |
| 40 | E11.3591 | Type 2 diabetes mellitus with other diabetic ophthalmic complication, right eye |
| 41 | E11.3592 | Type 2 diabetes mellitus with other diabetic ophthalmic complication, left eye |
| 42 | E11.3593 | Type 2 diabetes mellitus with other diabetic ophthalmic complication, bilateral |
| 43 | E11.3599 | Type 2 diabetes mellitus with other diabetic ophthalmic complication, unspecified eye |
| 44 | E11.36 | Type 2 diabetes mellitus with diabetic cataract |
| 45 | E11.39 | Type 2 diabetes mellitus with other diabetic ophthalmic complication |
| 46 | E11.40 | Type 2 diabetes mellitus with diabetic neuropathy, unspecified |
| 47 | E11.41 | Type 2 diabetes mellitus with diabetic mononeuropathy |
| 48 | E11.42 | Type 2 diabetes mellitus with diabetic polyneuropathy |
| 49 | E11.43 | Type 2 diabetes mellitus with diabetic autonomic neuropathy |
| 50 | E11.49 | Type 2 diabetes mellitus with other diabetic neurological complication |
| 51 | E11.51 | Type 2 diabetes mellitus with diabetic peripheral angiopathy without gangrene |
| 52 | E11.52 | Type 2 diabetes mellitus with diabetic peripheral angiopathy with gangrene |
| 53 | E11.59 | Type 2 diabetes mellitus with other circulatory complications |
| 54 | E11.610 | Type 2 diabetes mellitus with diabetic neuropathic arthropathy |
| 55 | E11.618 | Type 2 diabetes mellitus with other diabetic arthropathy |
| 56 | E11.621 | Type 2 diabetes mellitus with foot ulcer |
| 57 | E11.622 | Type 2 diabetes mellitus with other skin ulcer |
| 58 | E11.628 | Type 2 diabetes mellitus with other skin complications |
| 59 | E11.649 | Type 2 diabetes mellitus with hypoglycemia without coma |
| 60 | E11.65 | Type 2 diabetes mellitus with hyperglycemia |
| 61 | E11.69 | Type 2 diabetes mellitus with other specified complication |
| 62 | E11.8 | Type 2 diabetes mellitus with unspecified complications |
| 63 | E11.9 | Type 2 diabetes mellitus without complications |
| 64 | E13.9 | Other specified diabetes mellitus without complications |
| 65 | E13.10 | Other specified diabetes mellitus with ketoacidosis without coma |
| 66 | E13.22 | Other specified diabetes mellitus with diabetic chronic kidney disease |
| 67 | E13.29 | Other specified diabetes mellitus with other diabetic kidney complication |
| 68 | E13.3559 | Other specified diabetes mellitus with proliferative diabetic retinopathy with macular edema, unspecified eye |
| 69 | E13.42 | Other specified diabetes mellitus with diabetic polyneuropathy |
| 70 | E13.43 | Other specified diabetes mellitus with diabetic autonomic neuropathy |
| 71 | E13.49 | Other specified diabetes mellitus with other diabetic neurological complication |
| 72 | E13.620 | Other specified diabetes mellitus with diabetic dermatitis |
| 73 | E13.621 | Other specified diabetes mellitus with foot ulcer |
| 74 | E13.628 | Other specified diabetes mellitus with other skin complications |
| 75 | E13.65 | Other specified diabetes mellitus with hyperglycemia |

### S3 Appendix. List of ICD-10 Codes for Cardiovascular Disease Diagnosis

| Number | ICD Code | Description |
| --- | --- | --- |
| 1 | I25.10 | Atherosclerotic heart disease of native coronary artery without angina pectoris |
| 2 | I50.9 | Heart failure, unspecified |
| 3 | I63.9 | Cerebral infarction, unspecified |
| 4 | I65.23 | Occlusion and stenosis of bilateral carotid arteries |
| 5 | I65.29 | Occlusion and stenosis of unspecified carotid artery |
| 6 | I67.2 | Cerebral atherosclerosis |
| 7 | I67.9 | Cerebrovascular disease, unspecified |
| 8 | I73.9 | Peripheral vascular disease, unspecified |

### S4 Appendix. Methodological framework

Figure 5 provides a visual overview of the methodological framework of our study.

**Fig 5.**
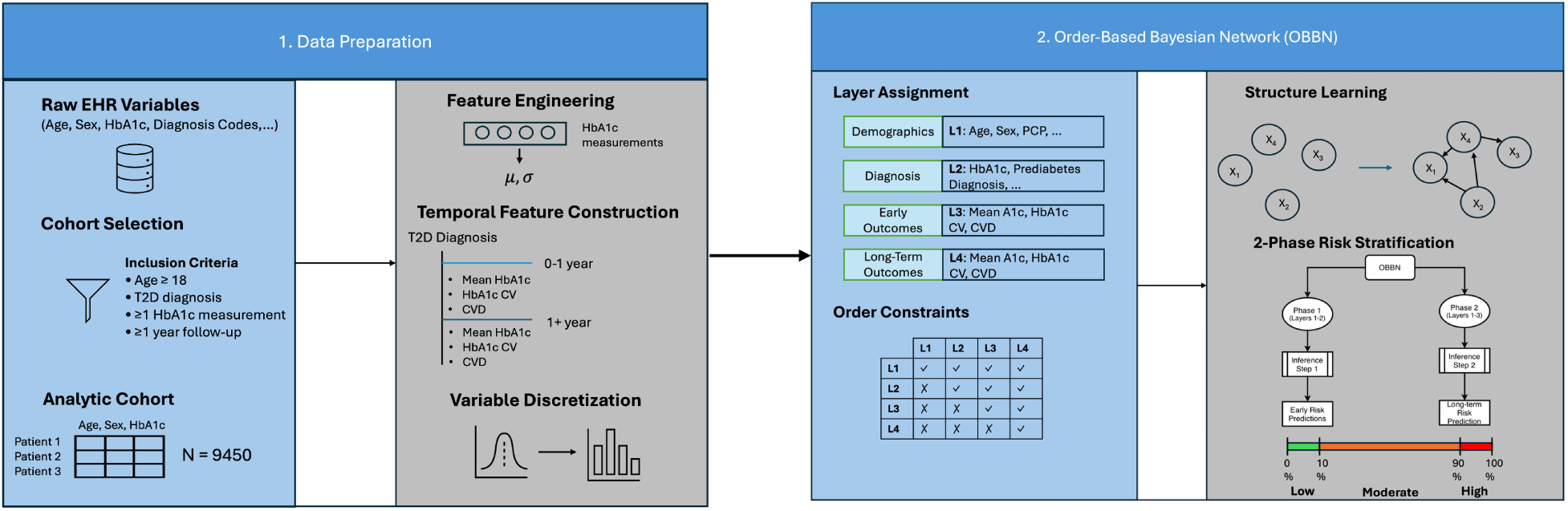
Overview of the methodological framework. (1) Data preparation: Raw EHR variables were filtered using cohort inclusion criteria to define the analytic cohort. Feature engineering and temporal feature construction were applied to derive early (0–1 year) and later (¿1 year) clinical outcomes, and continuous variables were discretized into categorical states for modeling. (2) Order-based Bayesian network (OBBN): Variables were assigned to temporally ordered layers representing demographics, diagnostic-stage factors, early outcomes, and long-term outcomes. Structure learning was performed under order constraints that restrict edges to flow from earlier to later layers. The trained OBBN was then used for two-phase risk stratification, where Phase 1 estimates early outcome risks using baseline variables (Layers 1–2), and Phase 2 estimates long-term risks using baseline and early outcomes (Layers 1–3). Predicted probabilities were stratified into low (≤ 10th percentile), moderate, and high (≥ 90th percentile) risk groups.

### S5 Appendix. Study Population Characteristics

**Table 7.**
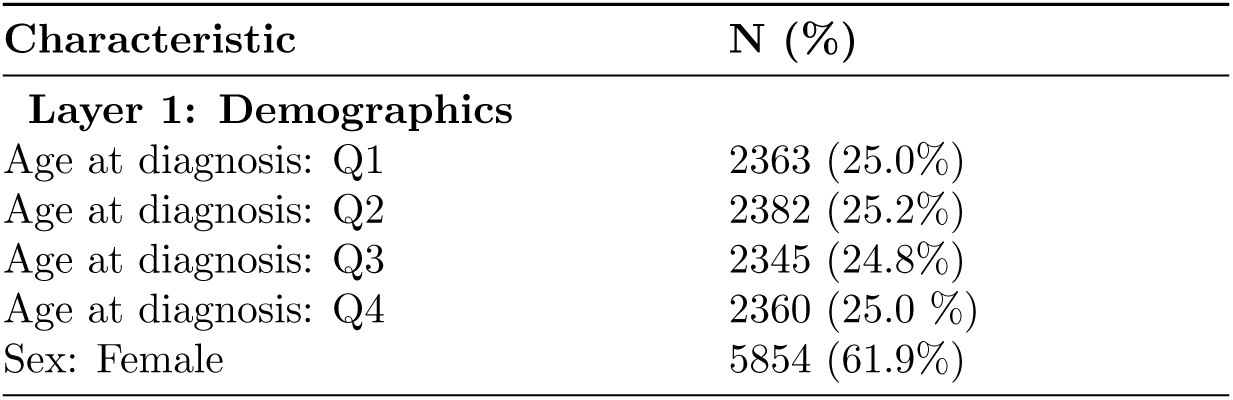
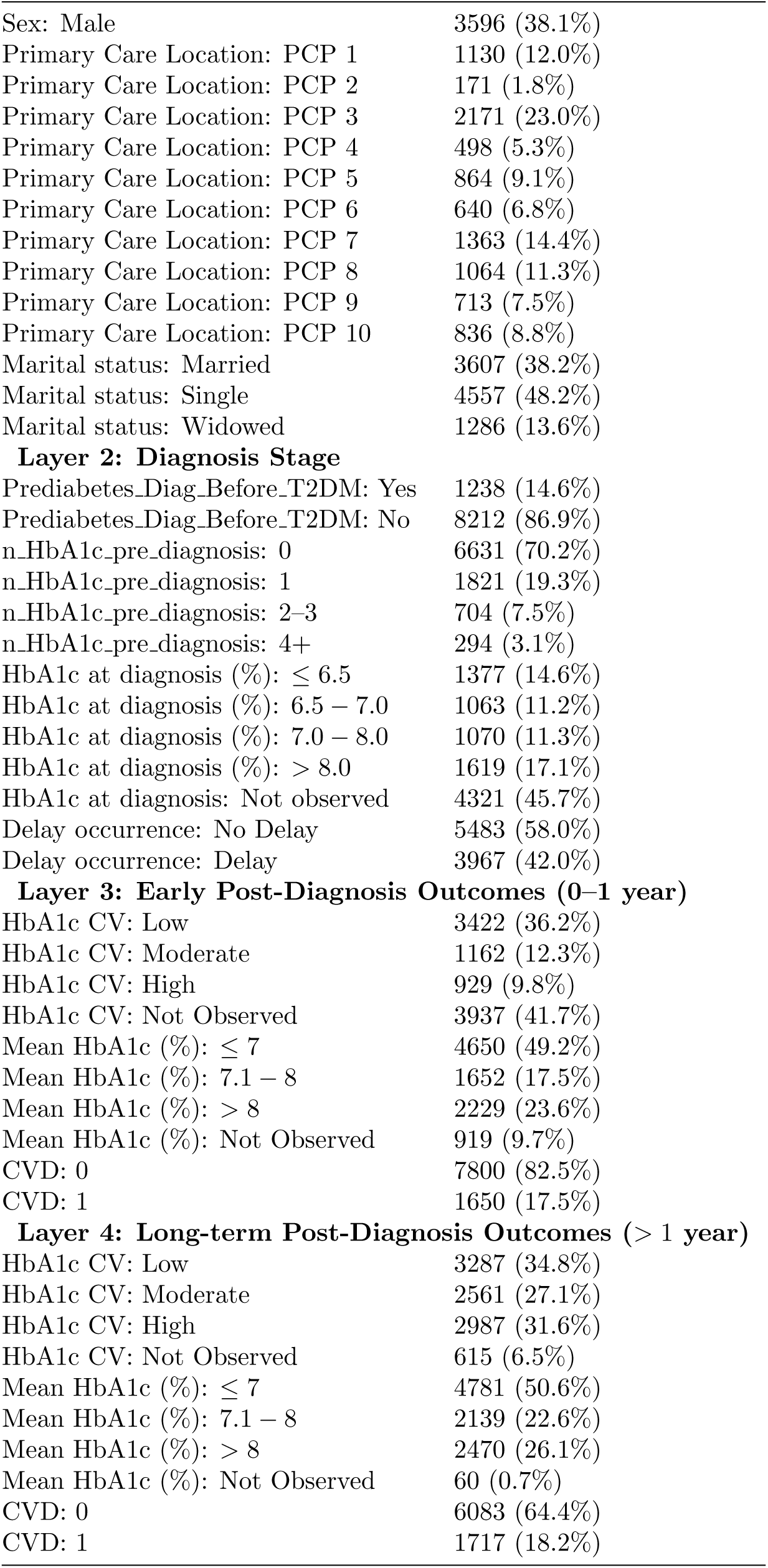
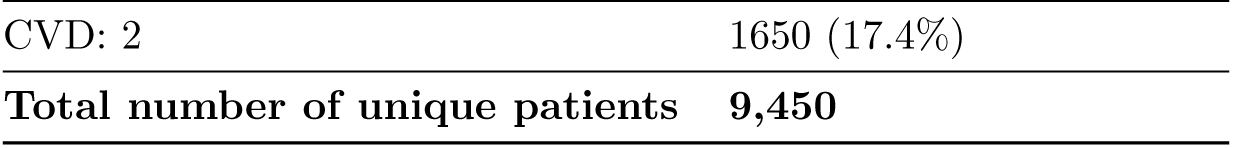
Study Population Characteristics. Categorical variables are reported as n (%). PCP stands for primary care provider, T2D for Type 2 Diabetes, and CV for coefficient of variation.

